# Molecular landscape and risk stratification in acute myeloid leukemia – insights from the real-world REFORM-AML cohort

**DOI:** 10.64898/2026.08.27.26361552

**Authors:** Daniel Tuyet Kristensen, Rasmus Froberg Brøndum, Michael Knudsen, Lykke Grubach, Claus Marcher, Birgitte Preiss, Maria Lyng Bibi, Estrid Høgdall, Tim Svenstrup Poulsen, Vibe Skov, Andreas Due Ørskov, Kirsten Grønbæk, Jakob Werner Hansen, Claudia Schöllkopf, Jack Bernard Cowland, Mette Klarskov, Marianne Tang Severinsen, Christopher Vejgaard, Ole Halfdan Larsen, Søren Vang, Martin Bøgsted, Anne Stidsholt Roug

## Abstract

Large genomically annotated acute myeloid leukaemia (AML) datasets exist, but population-based contemporary cohorts remain scarce. Here we report clinicopathological, genomic, and outcome data from Danish AML patients.

2,512 AML patients were identified between 2015-2022, of whom 33.8% had available NGS data (NGS+). In patients ≤70 years, baseline characteristics and outcomes were comparable between NGS+ and NGS− groups. In patients >70 years, more NGS+ patients received intensive treatment, but survival was similar among intensively treated patients. The distribution of mutations varied significantly by age and sex, with older age and male sex exhibiting higher frequencies of adverse-risk gene mutations.

In intensively treated NGS+ patients, ELN2017 stratified 5-year OS: 58.4% (favorable), 43.4% (intermediate), and 28.2% (adverse), with hazard ratios (HRs) of 0.63 (favorable) and 1.45 (adverse) relative to intermediate. ELN2022 yielded corresponding OS rates of 56.9%, 51.8%, and 29.7%, with HRs of 0.78 and 1.86. The two models had comparable predictive performance for OS in a time-dependent model.

In conclusion, outcomes of intensively treated AML patients were comparable irrespective of NGS status, underscoring the representativeness of the REFORM-AML database for the Danish AML population. Age and male sex correlated with adverse-risk mutations, and both ELN2017 and ELN2022 robustly predicted survival.

## INTRODUCTION

Acute myeloid leukemia (AML) is an aggressive hematological neoplasm and somatic mutations or cytogenetic alterations are found in more than 95% of cases with significant heterogeneity observed among patients[1,2]. Genetic alterations play a key role in the classification, risk stratification, and clinical decision-making[3–5]. Traditionally, AML has been defined by the presence of ≥20% blasts in the bone marrow or blood. In the latest International Consensus Classification (ICC), however, a new subgroup termed MDS/AML was introduced, defined by 10-19% blasts[3]. This change reflects the biological continuum and overlapping genetic alterations seen in myelodysplastic syndrome (MDS) with ≥10% blasts and AML offering a more meaningful basis for classification than the arbitrary 20% blast threshold[6].

The 2022 European LeukemiaNet (ELN2022) guideline on the diagnosis and classification of AML has replaced the previous 2017 version (ELN2017)[5,7]. Key updates include the assignment of *FLT3*-ITD mutated cases into the intermediate-risk group, the categorization and expansion of the list of myelodysplasia-related mutations, and new cytogenetic aberrations added to the adverse risk category. For the favorable risk group, the main change was the restriction of the favorable risk of previously biallelic *CEBPA* mutations to only include bZIP in-frame *CEBPA* mutations irrespective of allelic state[5,7].

Several large datasets and databases with genomically annotated AML cases have been collected since the gradual implementation of molecular testing[2,8–14]. However, challenges persist due to the limited availability of data for representative real-world cohorts including elderly patients, patients treated outside of clinical trials, and patients treated in a fully accessible healthcare system. These barriers can be addressed by establishing national population-based registries of genomic data. Population-based sequencing provides a comprehensive view of the genetic landscape of AML across different demographics, geographic regions, and environmental and social exposures and provides valuable epidemiological insights.

Here, we report on the establishment of the REFORM-AML database, a Danish national initiative designed to collect nationwide population-based panel sequencing data from patients with AML. In this real-world cohort, we describe the internal validity, characterize the genomic landscape and clinicopathological characteristics according to age and sex. Furthermore, we aimed to assess the prognostic performance of the ELN2022 compared with ELN2017 in unselected patients treated with intensive chemotherapy, focusing on each model to discriminate between risk categories and to predict survival at defined timepoints.

## MATERIALS AND METHODS

### Setting, data sources and study population

The study was conducted as a Danish nationwide population-based cohort study. The main data sources were the Danish Acute Leukemia Registry (DNLR) and The Danish Myelodysplastic Syndromes Database (DMDSD), both of which provide high coverage and data quality (see supplementary)[15,16]. The patient population consisted of cases with AML (≥20% blasts) reported to the DNLR and cases with MDS/AML defined as 10-19% blasts reported to the DMDSD, diagnosed between August 11, 2015, and September 21, 2022. Local pathological and genetic departments collected NGS files when available (NGS+) forming the REFORM-AML cohort (**Figure S1**). Patients from the same period without NGS data (NGS−) were included to analyze the internal validity of the NGS cohort to the general Danish AML and MDS/AML population. Patients treated with intensive chemotherapy and with available cytogenetic and NGS+ data were included to validate the prognostic significance of ELN2022 compared to ELN2017.

### Clinical information

Clinicopathological, treatment, and outcome data were obtained from the DNLR and the Danish DMDSD. Treatment was categorized as intensive, non-intensive, or palliative according to regimen intensity. Comorbidity was assessed using a modified Charlson Comorbidity Index[17,18], and disease ontogeny and cytogenetic risk were classified using established criteria (see Supplementary).

### Bioinformatical pipeline and variant calls

Leukemia-associated variants were assessed using center-specific targeted NGS panels (**Figure S2**) on pre-treatment bone marrow or peripheral blood samples obtained within 30 days of diagnosis. Sequencing data were reprocessed using standardized bioinformatic pipelines to harmonize variant calling across centers (see Supplementary).

### Statistical analysis

Baseline characteristics for the entire cohort were stratified by age (≤70 and >70 years) and by NGS+ and NGS−. Baseline characteristics for the cohort used to assess the prognostic performance of the ELN2022, stratification on risk category was presented. Categorical variables were presented as frequencies and percentages and continuous variables as median and interquartile range (IQR). Univariate comparison between strata of age were tested using a χ^2^-test for categorical variables and Mann-Whitney *U* test for continuous variables. The interactions between gene mutations, gene groups, and cytogenetic aberrations were investigated using a somatic interaction plot from the R-package *maftools*[19]. To test for significant interaction, the Fisher’s exact test was used while controlling the false discovery rate by the method of Benjamini and Hochberg[20]. The discriminative performance of the ELN2017 and the ELN2022 risk scores was assessed using univariable and multivariable Cox proportional hazard models, and estimated median OS, 2-year OS and 5-year OS from the Kaplan-Meier estimator, all with 95% confidence intervals (95% CI). The predictive performance for OS of ELN2017 and ELN2022 was compared with time-dependent receiver operating characteristic (ROC) curves using the R package *timeROC*[21]. All statistical analyses were done using R (version 4.4.2[22]).

## RESULTS

### The REFORM-AML cohort

During the study period, 2,512 patients were identified, including 2,203 with AML and 309 with MDS/AML. In total, 848 (33.8%, *n* = 101 for MDS/AML) patients were NGS+, and panel sequencing was more often performed in patients ≤70 years compared to >70 years (40.7% vs 29.1%) (**Table S1)**. For all samples, the median time from diagnosis to NGS sample date was 0 days (IQR, ±3 days).

### Comparison of NGS+ and NGS−

To investigate if NGS+ patients differed significantly from NGS− patients, a comparison was done within strata of age ≤70 years and >70 years. Baseline characteristics and univariate comparison can be seen in **Table S2**. In patients ≤70 years, no significant differences were observed except for diagnostic period and whether cytogenetics had been performed (both *p* < 0.001, **Table S2**). In patients >70 years, a similar pattern was seen with respect to diagnostic period and cytogenetics. Additionally, NGS+ patients were younger, had lower peripheral blast percentage, lower peripheral platelet count, and lower lactate dehydrogenase level (all *p* < 0.05). Treatment intensity also differed with more patients in the NGS+ strata allocated for intensive therapy and HSCT compared to NGS− (18.6% vs 9.7% and 2.5% vs 0.9%, respectively) (**Table S2**). No differences were seen in the comorbidity burden (**Table S2**). The crude median OS for NGS+ patients ≤70 years was 23.6 months (95% CI, 18.2-33.9) compared with 20.2 months (95% CI, 15.7-23.4) for NGS− (*p* = 0.22, **Figure 1A**). Among patients >70 years, the crude median OS was 6.5 months (95% CI, 5.5-8.6) for NGS+ compared with 3.5 months (95% CI, 3.0-4.0) for NGS− (*p <* 0.0001, Figure 1A). When restricting to patients treated with intensive chemotherapy (*n* = 906), the estimated crude median OS in patients ≤70 years was 56.5 months (95% CI, 31.2-NR) for NGS+ compared with 37.4 months (95% CI, 27.3-68.6) for NGS− (*p* = 0.58, **Figure 1B**). Similarly, for age >70 years, the median OS was estimated to 21.1 months (95% CI, 15.2-33.8) for NGS+ compared with 17.2 months (95% CI, 12.6-22.0) for NGS− (*p* = 0.17, **Figure 1B**).

**Figure 1:**
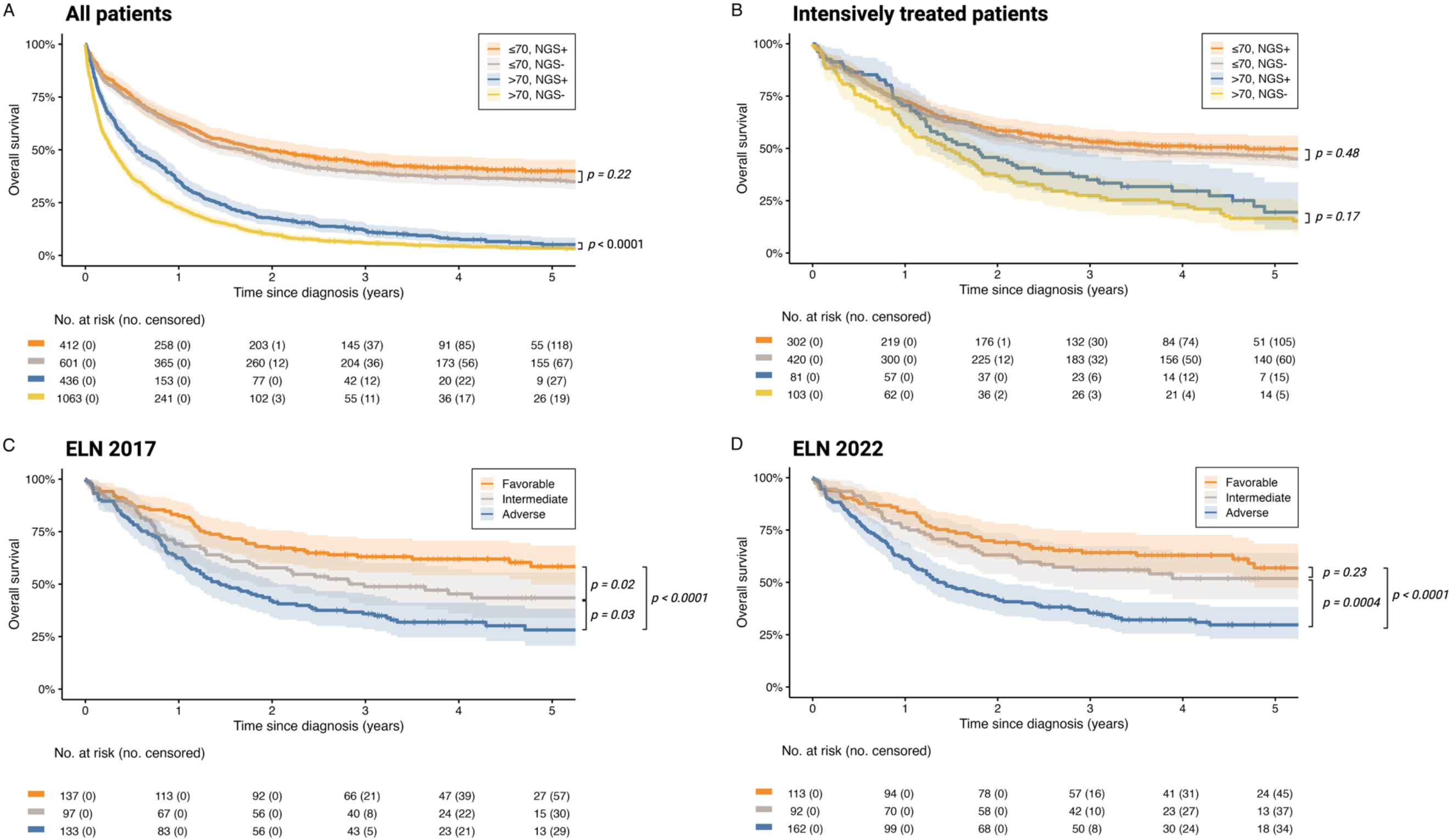
Crude overall survival for A) All patients stratified by NGS+/NGS− and age ≤70 and >70 years (*n* = 2,512), B) All patients intensively treated stratified by NGS+/NGS− and age ≤70 and >70 years (*n* = 906), C) Intensively treated patients with NGS and cytogenetic information stratified according to ELN2017 (*n* = 367), and D) Intensively treated patients with NGS and cytogenetic information stratified according to ELN2022 (*n* = 367). *P*-values from log-rank test.

### Real-world mutational landscape

The distribution of somatic mutations according to sex, age strata, and AML vs MDS/AML from the NGS+ cohort with available cytogenetics (*n* = 785) can be seen in **Figure 2A-D** and co-mutational patterns in **Figure 3A-B and S3A-D**. Patients ≥70 years had higher frequencies of mutations in *RUNX1* and *TET2*, whereas younger patients had higher frequencies of *DNMT3A*, *FLT3* (including TKD and ITD variants) and bZIP in-frame *CEBPA* mutations (**Figure 2B**). The genetic landscape also varied according to sex, where male patients had higher proportions of mutations in adverse risk genes including *RUNX1*, *SRSF2*, *ASXL1, ZRSR2,* and *U2AF1*, whereas female patients had higher proportions of *DNMT3A* and *NPM1* (**Figure 2C**). Mutations in *NPM1,* in frame bZIP *CEBPA*, and *FLT3* (including TKD and ITD variants) were almost exclusively seen in AML and only sporadically in MDS/AML. Also, *DNMT3A* and *IDH1* mutations were more frequent in AML compared to MDS/AML. In contrast, mutations in *TP53* and *SRSF2* were more commonly found in MDS/AML compared with AML (**Figure 2D**).

**Figure 2:**
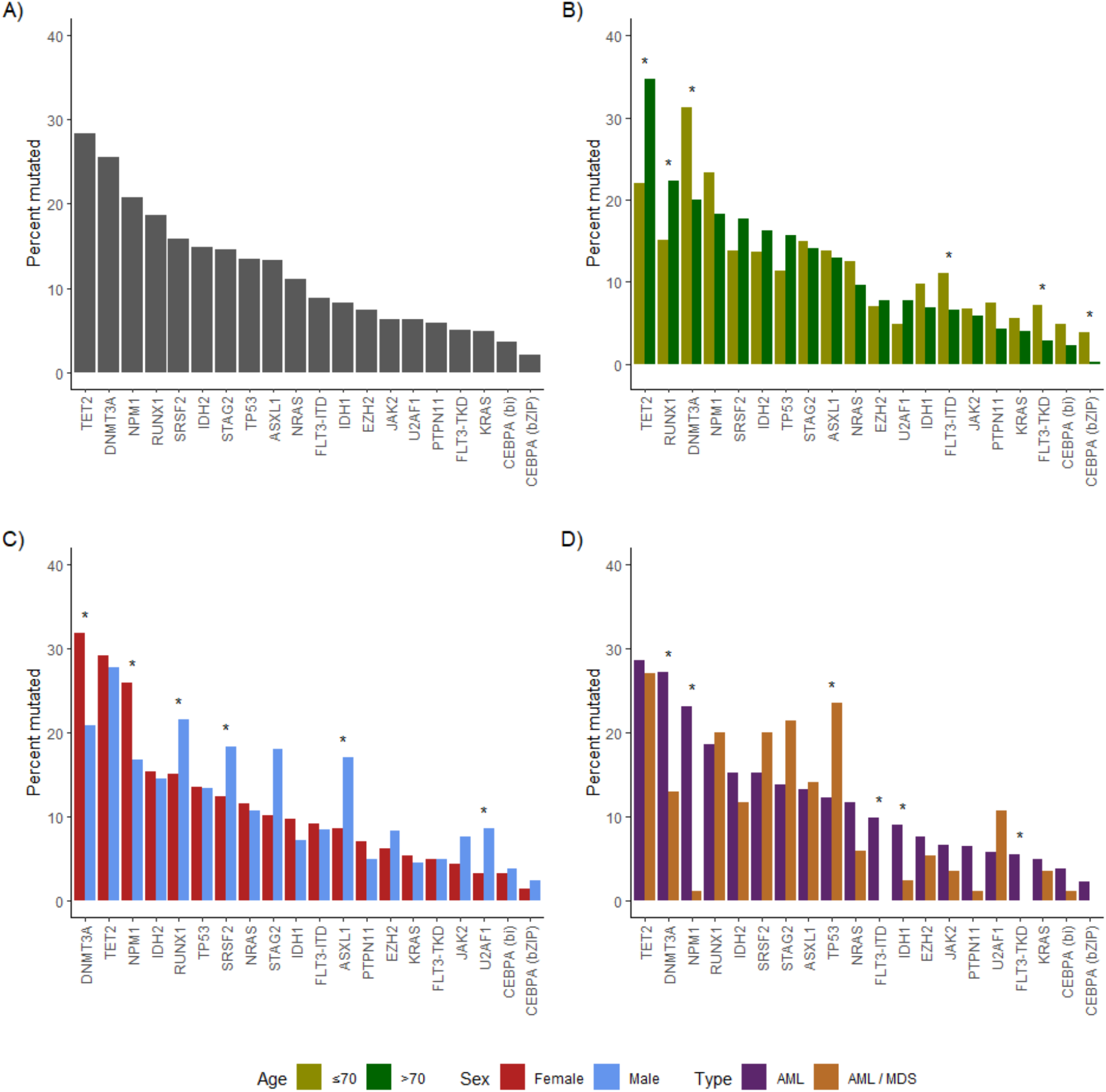
Barplot showing the distribution of selected genes in A) All patients (*n* = 785), B) For age ≤70 year (*n* = 390) and >70 years (*n* = 395), C) For male (*n* = 446) and female (*n* = 339), and D) For AML (*n* = 700) and MDS/AML (*n* = 85). * Denotes p<0.05 (Fisher’s exact test).

**Figure 3:**
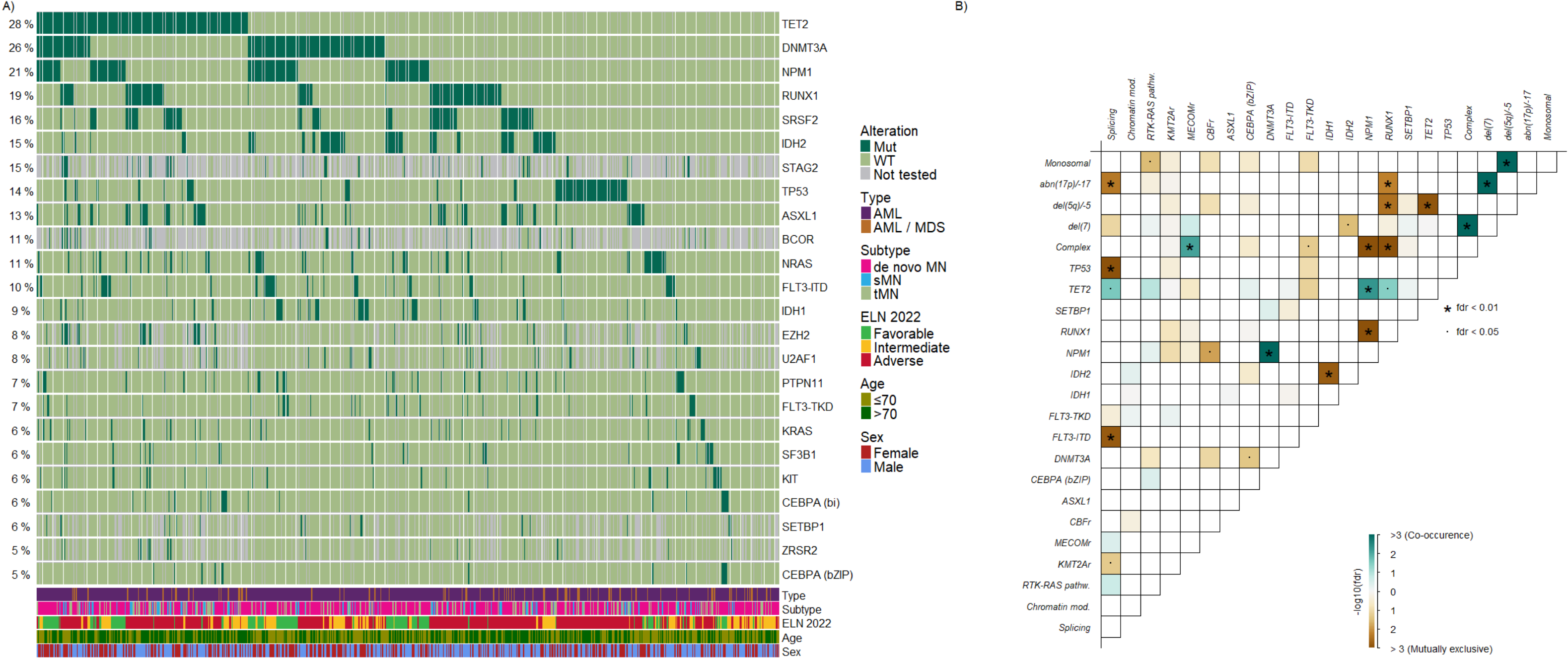
The mutational landscape of the cohort (*n* = 785), A) Oncoplot displaying mutations and selected clinical characteristics for the cohort, and B) Mutational interaction plot showing the interaction between single genes, group of genes, and cytogenetic aberrations, deeper brown color denotes mutually exclusive interaction and deeper green color denoted co-occurrence, fdr denotes the false discovery rate corrected *p*-value. Chromatin mod. denotes genes involved in chromatin modification (*BCOR*, *EZH2*, *STAG2*), Spicing denotes genes involved in RNA splicing (*SF3B1*, *SRSF2*, *ZRSR2*, *U2AF1*), RTK-RAS-pathw. denotes genes involved in RTK-RAS-pathway (*KIT*, *HRAS*, *KRAS*, *NRAS*, *PTPN11*, *CBL*, *BRAF*, and *NF1*). CBFr denotes the core-binding factor defining alterations inv(16)/t(16;16) and t(8;21)(q22;q22), *KMT2A*r denotes rearranged *KMT2A,* t(v;11q23), and *MECOM*r denotes *MECOM*-rearrangements, t(3q26.2;v). Complex karyotype is defined as three or more unrelated chromosome abnormalities in the absence of other class-defining recurring genetic abnormalities and hyperdiploid karyotype. Abbreviations: CK, complex karyotype; MK, monosomal karyotype.

The interaction between mutations in individual genes, gene groups, and cytogenetic aberrations and distribution according to clinical characteristics are given in **Figure 3A-B** and **Figure S3**. ELN2022 adverse alterations were found to cluster together, including mutated *TP53*, del(5q), −5, −7, −17/abn(17p), monosomal and complex karyotypes, whereas these were mutually exclusive from more favorable risk alterations, e.g., mutated *NPM1* (**Figure 3B**). Mutated *NPM1* was found to co-occur with *FLT3*, *DNMT3A,* and *TET2*, whereas mutated *IDH1* or *IDH2* were mutually exclusive (**Figure 3B**).

### European LeukemiaNet risk stratification

To assess the prognostic significance of the ELN2017 and ELN2022, we analyzed a subset of 367 NGS+ patients treated with intensive chemotherapy, comprising 354 (96.5%) with AML and 13 (3.5%) with MDS/AML. Baseline characteristics for this cohort stratified on ELN2022 risk category are displayed in **Table 1**.

**TABLE 1.** Selected baseline characteristics of the cohort used to validate ELN2022.

| Variable | ELN2022 risk category |  |  |  |
| --- | --- | --- | --- | --- |
|  | All | Favorable | Intermediate | Adverse |
| **Numbers**, proportion of all (%) | 367 (100) | 113 (30.8) | 92 (25.1) | 162 (44.1) |
| **Sex**, male, *n* (%) | 190 (51.8) | 54 (47.8) | 43 (46.7) | 93 (57.4) |
| **Age**, median, years (IQR) | 63 (51-69) | 58 (49-69) | 59 (49-66) | 65 (59-70) |
| **Age >70 years**, *n* (%) | 75 (20.4) | 21 (18.6) | 14 (15.2) | 40 (24.7) |
| **Comorbidity index**, *n* (%) |  |  |  |  |
| ≤2 | 215 (58.6) | 73 (64.6) | 59 (64.1) | 83 (51.2) |
| 3-5 | 133 (36.2) | 32 (28.3) | 30 (32.6) | 71 (43.8) |
| ≥6 | 19 (5.2) | 8 (7.1) | 3 (3.3) | 8 (4.9) |
| **Diagnostic period**, *n* (%) |  |  |  |  |
| 2015-2017 | 47 (12.8) | 13 (11.5) | 10 (10.9) | 24 (14.8) |
| 2018-2020 | 217 (59.1) | 71 (62.8) | 55 (59.8) | 91 (56.2) |
| 2021-2022 | 103 (28.1) | 29 (25.7) | 27 (29.3) | 47 (29.0) |
| **Disease ontogeny\***, *n* (%) |  |  |  |  |
| *De novo* | 295 (80.4) | 101 (89.4) | 73 (79.3) | 121 (74.7) |
| Antecedent hematological disease | 23 (6.3) | 1 (0.9) | 3 (3.3) | 19 (11.7) |
| Therapy-related | 49 (13.4) | 11 (9.7) | 16 (17.4) | 22 (13.6) |
| **BM blast**, median %, (IQR) | 57 (35-80) | 66 (41-80) | 65 (45-85) | 46 (26-71) |
| **BM blast <20%**, *n* (%) | 13 (3.5) | 0 (0.0) | 2 (2.2) | 11 (6.8) |
| **PB blast**, median %, (IQR) | 22 (4-55) | 26 (9-60) | 30 (5-62) | 17 (2-49) |
| **Platelet**, median, ×10<sup>9</sup>/L, (IQR) | 66 (33-122) | 56 (29-123) | 85 (36-150) | 60 (33-107) |
| **WBC**, median, ×10<sup>9</sup>/L, (IQR) | 9.4 (2.6-40.2) | 16.4 (5.5-51.4) | 8.5 (1.9-58.6) | 5.8 (2.2-31.1) |
| **LDH**, median, U/L (IQR) | 376 (237-650) | 402 (284-686) | 396 (215-921) | 337 (224-518) |
| **Cytogenetic risk<sup>#</sup>**, *n* (%) |  |  |  |  |
| Favorable | 21 (5.7) | 21 (18.6) | 0 (0.0) | 0 (0.0) |
| Intermediate | 278 (75.7) | 92 (81.4) | 92 (100.0) | 94 (58.0) |
| Adverse | 68 (18.5) | 0 (0.0) | 0 (0.0) | 68 (42.0) |
| **ELN2017 risk**, *n* (%) |  |  |  |  |
| Favorable | 137 (37.3) | 112 (99.1) | 17 (18.5) | 8 (4.9) |
| Intermediate | 97 (26.4) | 0 (0.0) | 69 (75.0) | 28 (17.3) |
| Adverse | 133 (36.2) | 1 (0.9) | 6 (6.5) | 126 (77.8) |
| **HSCT**, *n* (%) | 140 (38.1) | 32 (28.3) | 42 (45.7) | 66 (40.7) |
| Abbreviations: BM, bone marrow; ELN, European LeukemiaNet; HSCT, hematopoietic stem cell transplantation; IQR, interquartile range (25th to 75th percentiles); LDH, lactate dehydrogenase; PB, peripheral blood; WBC, white blood cell count. *Antecedent hematological disease takes precedence over therapy-related <sup>#</sup>Risk according to the European LeukemiaNet 2022 risk score[5]. |  |  |  |  |

Using ELN2017, 37.3%, 26.4%, and 36.2% of patients were categorized as favorable, intermediate, and adverse risk, respectively. A total of 60/367 (16.3%) changed risk category when moving from ELN2017 to ELN2022, where most cases were reclassified from intermediate to adverse (28/60, 44.7%), followed by favorable to intermediate (17/60, 28.3%) and favorable to adverse (8/60, 13.3%) (**Figure 4A**). In ELN2022, 30.8% were categorized as favorable, 25.1% as intermediate, and 44.1% as adverse. The distribution of risk categories according to ELN2017 and ELN2022 across age and sex strata is shown in **Figure 4B**. In ELN2017, male patients had a numerically higher proportion of adverse risk, and a lower proportion of favorable risk compared with female patients, with a similar trend observed across age strata. This pattern persisted in ELN2022, where the skewed distribution of adverse vs favorable risk was even more pronounced in males and patients aged >70 years, compared with female sex and age ≤70 years (**Figure 4B**). All these differences were numerical but not statistically significant (χ^2^-test *p* >0.05).

**Figure 4:**
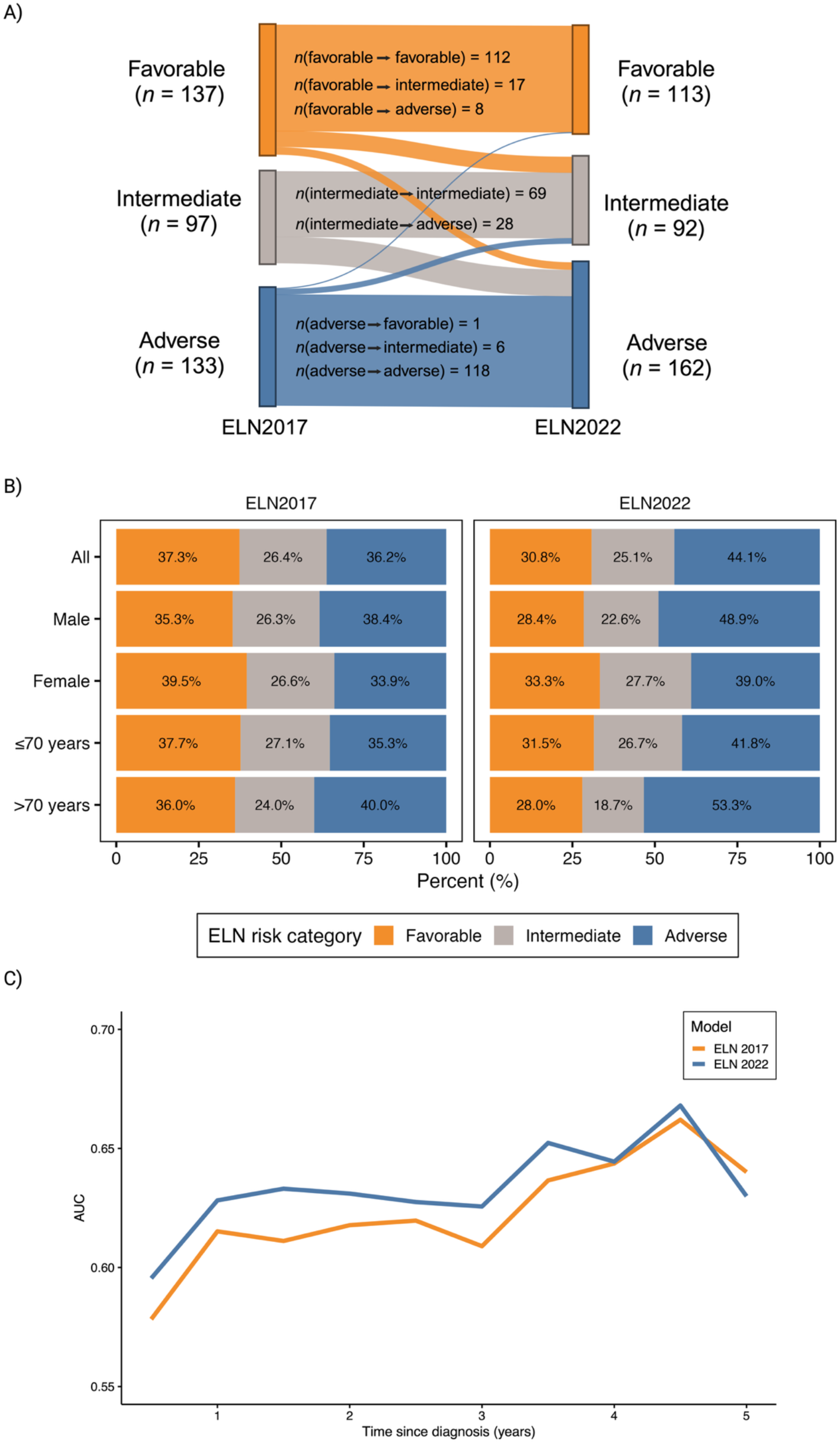
A) Sankey plot showing the reclassification of the cohort from ELN2017 to ELN2022, B) Distribution of risk classification of ELN2017 and ELN2022 among all patients, strata of age, and strata of sex (no significant differences were observed within strata using χ^2^-test), and C) Plot of time-varying receiver operator curves for ELN2017 and ELN2022.

### Comparison of overall survival and prognostic performance of ELN2017 and ELN2022

For the 367 NGS+ patients treated with intensive chemotherapy and with a median follow up of 51.1 months (IQR, 38.4-68.1), the median OS was 35.0 months (95% CI, 25.9-57.3), and the median OS for risk categories of ELN2017 and ELN2022 are given in **Table 2**. For ELN2017, crude 5-year OS discriminated between all risk strata with a ∼15% difference in OS between favorable and intermediate, as well as between intermediate and adverse risk groups. The 5-year OS for favorable risk was 58.4% (95% CI, 49.8-68.3) compared to 43.4% (95% CI, 33.9-55.5) and 28.2% (95% CI, 20.7-38.3) for intermediate and adverse risk, respectively (**Table 2**, **Figure 3C**). Compared to the intermediate risk group, the crude hazard ratio (HR) for death was 0.63 (95% CI, 0.43-0.92) for favorable and 1.45 (95% CI, 1.04-2.03) for adverse risk. When controlling for age and sex, the estimates remained largely unchanged; however, the discrimination between intermediate and adverse was no longer statistically significant (**Table 2**).

**TABLE 2:**
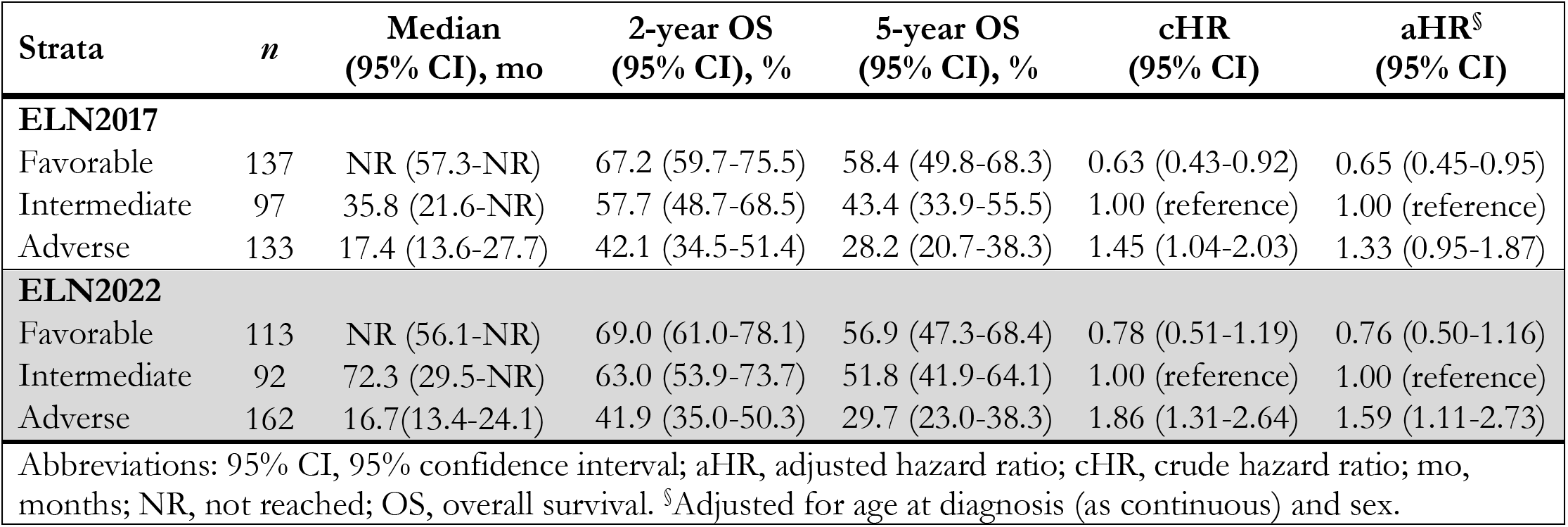
Median OS, crude and adjusted hazard ratios (HRs) for OS from univariate and multivariate cox proportional hazard models by risk category from ELN2017 and ELN2022.

| Strata | *n* | Median<br>(95% CI), mo | 2-year OS<br>(95% CI), % | 5-year OS<br>(95% CI), % | cHR<br>(95% CI) | aHR<sup>§</sup><br>(95% CI) |
| --- | --- | --- | --- | --- | --- | --- |
| **ELN2017** |  |  |  |  |  |  |
| Favorable | 137 | NR (57.3-NR) | 67.2 (59.7-75.5) | 58.4 (49.8-68.3) | 0.63 (0.43-0.92) | 0.65 (0.45-0.95) |
| Intermediate | 97 | 35.8 (21.6-NR) | 57.7 (48.7-68.5) | 43.4 (33.9-55.5) | 1.00 (reference) | 1.00 (reference) |
| Adverse | 133 | 17.4 (13.6-27.7) | 42.1 (34.5-51.4) | 28.2 (20.7-38.3) | 1.45 (1.04-2.03) | 1.33 (0.95-1.87) |
| **ELN2022** |  |  |  |  |  |  |
| Favorable | 113 | NR (56.1-NR) | 69.0 (61.0-78.1) | 56.9 (47.3-68.4) | 0.78 (0.51-1.19) | 0.76 (0.50-1.16) |
| Intermediate | 92 | 72.3 (29.5-NR) | 63.0 (53.9-73.7) | 51.8 (41.9-64.1) | 1.00 (reference) | 1.00 (reference) |
| Adverse | 162 | 16.7(13.4-24.1) | 41.9 (35.0-50.3) | 29.7 (23.0-38.3) | 1.86 (1.31-2.64) | 1.59 (1.11-2.73) |
| Abbreviations: 95% CI, 95% confidence interval; aHR, adjusted hazard ratio; cHR, crude hazard ratio; mo, months; NR, not reached; OS, overall survival. <sup>§</sup>Adjusted for age at diagnosis (as continuous) and sex. |  |  |  |  |  |  |

When stratifying patients according to ELN2022, the crude 5-year OS was 56.9% (95% CI, 47.3-68.4), 51.8% (95% CI, 41.9-64.1) and 29.7% (95% CI, 23.0-38.3) for favorable, intermediate and adverse risk, respectively (**Table 2**, **Figure 3D**). Here, the discrimination between favorable and intermediate risk was less pronounced compared to ELN2017, whereas the gap between intermediate and adverse was more pronounced. Using intermediate risk as reference, the estimated HR for death was 0.78 (95% CI, 0.51-1.19) for favorable risk and 1.86 (95% CI, 1.31-2.64) for adverse. Adjusting for age and sex had only limited influence on the estimates (**Table 2**). Consequently, ELN2022 effectively distinguished favorable and intermediate risk groups from the adverse risk group, whereas it did not show a significant distinction between favorable and intermediate risk groups.

The predictive performance of ELN2017 and ELN2022 for OS was evaluated using time-dependent ROC curves for OS at six months intervals by ranking the risk groups (**Figure 4C**). Although ELN2022, did not show statistically significant superiority over ELN2017 at any timepoint, it generally demonstrated a numerically higher area under the curve (AUC) within the first 3.5 years following diagnosis (**Figure 4B**).

## DISCUSSION

This study presents the population-based Danish REFORM-AML cohort and evaluates its representativeness and internal validity. NGS+ and NGS− patients had comparable characteristics among patients ≤70 years. In patients >70 years, NGS+ status was associated with a higher likelihood of receiving intensive chemotherapy, resulting in differences in OS that were likely driven by selection bias and treatment allocation, including the introduction of venetoclax-based regimens. Consistently, outcomes were similar between NGS+ and NGS− patients when analyses were restricted to intensively treated patients. Overall, the REFORM-AML cohort appears representative of a real-world, unselected AML population.

In AML, increasing age is associated with an adverse genetic risk with *NPM1* and *CEBPA* mutation frequencies decreasing, and mutations in *TP53*, *RUNX1*, and *ASXL1* becoming more frequent[23]. This was also observed in the REFORM-AML cohort, where patients >70 years more frequently harbored *RUNX1* mutations and were classified as adverse risk by both ELN2017 and ELN2022. Interestingly, we observed marked sex-based disparity in mutational patterns with males showing significantly higher frequencies of adverse-risk mutations (*RUNX1*, *SRSF2*, *ASXL1,* and *U2AF1)*, while females more often harbored the favorable *NPM1*-mutation, without a significant increase in *FLT3*-ITD. Sex-related differences in distribution of mutations have been examined in only a few previous studies. Ozga *et al.* investigated sex variations primarily in younger *de novo* AML patients enrolled in prospective clinical trials[24]. Consistent with our findings, they reported higher frequencies of *ASXL1*, *SRSF2*, *U2AF1*, *RUNX1*, and *KIT* mutations in males, while mutated *DNMT3A*, *NPM1,* and *WT1* were more common in females[24]. Furthermore, when analyzing mutations by functional groups, males had higher frequencies of mutations involved in chromatin remodeling, spliceosome, and myelodysplasia-related categories[24]. Similar findings have been reported in the German Münchner Leukämielabor cohort[25]. Collectively, current evidence indicates that male sex is associated with a more adverse genetic risk in AML. However, it remains unclear whether genetic aberrations fully account for the observed negative prognostic impact of male sex [26].

MDS/AML cases were included in the REFORM-AML cohort. Although sharing key features with AML≥20% blast, MDS/AML showed a distinct mutational profile. Male sex, age >70 years, and MDS/AML were associated with higher frequencies of adverse-risk mutations. This was also reflected when employing ELN2022, where male sex and elderly had higher frequencies of adverse risk profiles, a trend that was less pronounced in ELN2017. This likely reflects the expanded inclusion of myelodysplasia-related mutations in ELN2022, which are associated with older age and secondary AML [5][23,27,28].

To evaluate the prognostic performance of ELN2022 and ELN2017, we first assessed their ability to distinguish risk categories and then their accuracy in predicting survival at specific time points. First, 16% of patients were reclassified under ELN2022, with most shifting to a higher-risk category. This aligns with other studies with reclassification rates of 13-23%, predominantly to higher risk categories[14,29–34]. While ELN2017 effectively distinguished between favorable, intermediate, and adverse risk groups based on 5-year OS and HR for death, ELN2022 primarily separated favorable and intermediate from adverse risk, albeit with a higher HR. Previous studies have found diverging results: Two studies found better separation of risk groups using ELN2022[29,31] while four report similar or worse discrimination of ELN2022 compared to ELN2017[30,32–34]. OS prediction in the REFORM-AML cohort found no significant differences between ELN 2017 and ELN2022. Three other studies have compared the predictive performance between ELN2022 and ELN 2017, and also reported diverging results, ranging from improved prediction with ELN2022[33], to similar[14], or even inferior relative to ELN2017[34]. A key limitation of prior studies is their reliance on older clinical trial data, some dating back to the 1980s and 1990s, that do not reflect contemporary AML treatment. This is reflected in the 5-year OS estimates from these studies ranging from 48-53% for favorable, 22-32% for intermediate, and 7-16% for adverse risk groups, which are lower than those observed in our contemporary, non-selected, real-world population. These differences highlight the challenges of applying historical data in modern practice, as prognostic models evolve with treatment advances. A notable example is *FLT3*-ITD mutated AML. In ELN17, patients were classified based on *NPM1* co-mutation and *FLT3*-ITD allelic ratio. In ELN2022, the reclassification of *FLT3*-ITD without adverse cytogenetics as intermediate, reflects improved outcomes associated with *FLT3*-ITD targeted therapy[5,7]. The most comparable contemporary is the PETHEMA cohort[30], which includes realworld patients treated in modern era. Consistent with our findings, it reported fewer favorable and intermediate-risk cases, similar sex and age distribution, a reclassification rate of 14.5%, and comparable OS predictions for ELN2017 and ELN2022. However, PETHEMA’s global cohort may reflect differences in genomic testing and treatment practices.

The REFORM-AML cohort was generated using different NGS panels with variable gene coverage. Consequently, some ELN2022-defined myelodysplasia-related genes, including *STAG2* and *BCOR*, were not assessed in all patients, potentially introducing misclassification bias. Expanding the number of intensively treated patients with both cytogenetic and sequencing data would increase the statistical power to detect differences between risk groups. However, the real-world design of the cohort is a major strength, reflecting routine clinical practice. Importantly, all NGS results were available to treating clinicians and informing therapeutic decision-making.

The REFORM-AML cohort reflects contemporary AML management, with routine use of FLT3 inhibitors since 2018, addition of gemtuzumab ozogamicin to DA3+7 for favorable/intermediate-risk disease, and CPX-351 for sAML/tAML since 2019. Patients also benefited from standardized infection prophylaxis, modern intensive care, equal access to healthcare resources including HSCT, and the availability of venetoclax-based salvage therapies, making the cohort highly representative of current clinical practice [35,36].

In conclusion, analysis of the contemporary, population-based REFORM-AML cohort demonstrated that NGS+ patients were representative of the general Danish AML population, supporting the generalizability of the cohort. The comparison of ELN2017 and ELN2022 risk classifications revealed similar performance in predicting survival. As NGS becomes increasingly integrated into AML management, including in relapsed and refractory disease and in patients ineligible for intensive chemotherapy, molecular risk stratification is expected to play an even greater role in prognostication and treatment selection. It is therefore of critical importance that risk scoring systems reflect contemporary AML treatment, outcomes and diagnostic work up.

## Data Availability

Per Danish law, the data cannot be shared directly; however, they can be accessed through application. Additional information is available on request from the corresponding author.

## ACKNOWLEDGEMENT

The Danish Healthcare Quality Institute is acknowledged for providing and governing data from the Danish Acute Leukemia Registry (DNLR). Appreciation is expressed to those who contributed to the formation and data collection of the DNLR.

## ETHICAL APPROVAL AND PATIENT CENSENT STATEMENT

The study was approved and registered the North Denmark Region (ID: 2021-011009 and 2021-197). According to Danish health care legislation, patient consent has been waived according to the approval.

## AUTHOR CONTRIBUTIONS

**Conception and design**: Daniel Tuyet Kristensen, Rasmus Froberg Brøndum, Martin Bøgsted, and Anne Stidsholt Roug

**Provision of study materials or patients:** Daniel Tuyet Kristensen, Marianne Tang Severinsen, Lykke Grubach, Mette Klarskov Andersen, Vibe Skov, Birgitte Preiss, Maria Bibi Lyng, Estrid Høgdall, Tim Svenstrup Poulsen, Andreas Due Ørskov, Jakob Werner Hansen, Claudia Schöllkopf, Kirsten Grønbæk, Jack Bernard Cowland, Mette Klarskov Andersen, Claus Marcher, Christopher Vejgaard, and Anne Stidsholt Roug

**Collection, assembly and analysis of data**: Daniel Tuyet Kristensen, Rasmus Froberg Brøndum, Michael Knudsen, Ole Halfdan Larsen, Søren Vang, Martin Bøgsted, and Anne Stidsholt Roug.

**Data interpretation**: All authors

**Manuscript writing**: Daniel Tuyet Kristensen and Anne Stidsholt Roug

**Critical revision and final approval of manuscript:** All authors

**Accountable for all aspects of the work**: All authors

## Supplementary material

### Data source and definitions

#### The Danish Acute Leukemia Registry (DNLR) and Danish Myelodysplastic Syndromes Database (DMDSD)

A collection of hematologic registries exists under the governance of the Danish Society of Hematology and the Danish Healthcare Quality Institute. The registries are disease specific and include patients with different hematologic diagnoses.

For this study, the Danish Acute Leukemia Registry (DNLR) and Danish Myelodysplastic Syndromes Database (DMDSD), established in 2000 and 2010, respectively, were used to define a study population and retrieve data on the disease course. Both data sources include prospectively collected data on all cases of adult AML and MDS with the primary aim of improving quality of care and treatment through insights into epidemiological factors and identification of prognostic and predictive factors for outcome. All participating hospitals departments treating AML are obliged to report requested data and patient consent is not required. The physician registers new cases using standardized registrations forms found in an online registration-based system[1]. A validation study found an AML-coverage >99%, and for thirty variables obtained, the study found a completeness between 60-100 %, and a positive predictive value (PPV) ranging from 89-100% in the DNLR[2]. Similarly, the DMDSD offers high coverage of >95% after 2015 and the predictive values ranged between 64-100%, and for 36/48 variables the predictive value exceeded 90%[3].

#### Bioinformatic pipeline and variant call

For data produced using the Ion Torrent^TM^ sequencing technology, where the majority was Oncomine Myeloid Research Assay[4], BAM files were uploaded to an Ion Reporter server (Thermo Fisher Scientific Inc., Waltham, MA, USA), which was used to call and filter variants according to a proprietary pipeline and filters (Oncomine Extended filter chain, version 5.18). Reads were initially aligned to the hg19 reference genome and subsequently the resulting variant call format (VCF) files were lifted to the hg38 reference genome.

For Illumina sequencing, fastq files were re-processed using an in-house pipeline at Aarhus University’s high-performance computer, GenomeDK. Here, adapter sequences were trimmed using cutadapt, and the trimmed reads were mapped to the hg38 reference genome using BWA-MEM[5,6]. For samples sequenced with unique molecular identifiers (UMIs), consensus reads from reads with identical UMI sequences mapping to the same genomic positions were created using fgbio best practices, and for the remaining samples duplicates were marked using Picard MarkDuplicates[7,8]. Variants were called using GATK Mutect2 and filtered using GATK FilterMutectCalls[9,10]. To increase sensitivity, filtered variants outside repeat-masked regions were elevated to PASS if they were called by either VarScan or Pindel[10,11]. Finally, additional INDEL calls by Pindel in the *FLT3*-ITD region were added to the set of final variants. The final set of VCF files from both technologies were converted to MAF using *vcf2maf* which filters out known germline variants and adds functional annotation[12].

To combine individual samples into a table of variants for statistical analysis, MAF files were processed in R (version 4.4.2[13]) using *mafTools* v2.22.0[14]). In short, data from different panels were filtered to only keep variants from the respective target areas with a PASS filter value from vcf2maf, variant allele frequency (VAF) of at least 5%, or, alternatively, a VAF of at least 2% if the variant was included in the hotspot list from the ChromoSeq pipeline[15]. A subgroup of patients was not tested for all genes (genes defined as myelodysplasia-related, see Figure S2) in the ELN22, in this event, risk assignment by ELN22 was set for the available information and no available information was set as non-mutated.

#### Supplementary treatment information

For patients categorized as intensively treated, regimens comprised DA-like (daunorubicin [or another anthracycline/anthracycline-related compound] + cytarabine), DA-like + add on (DA-like plus etoposide, all-trans retinoic acid, gemtuzumab ozogamicin, clofarabine, or midostaurin), FLAG-Ida-like (fludarabine + cytarabine + idarubicin [or another anthracycline/anthracycline-related compound] + granulocyte colony-stimulating factor), or liposomal DA (CPX351) (see Table S1 for additional information). During the study period, double induction was standard clinical practice.

The treatment of acute leukemias including AML Denmark have in Denmark been dictated by national consensus guidelines provided by the Danish Acute Leukemia Group (ALG). These guidelines are updated every other year or earlier, if major changes occur (https://leukemia.dk). During the study period, the standard first line therapy has been “DA 3+10” following the British NCRI scheme[16], due to Denmark’s participation in the NCRI AML15, AML16, AML17, AML18, and latest AML19 protocols.

## Supplemental figures

**Figure S1:**
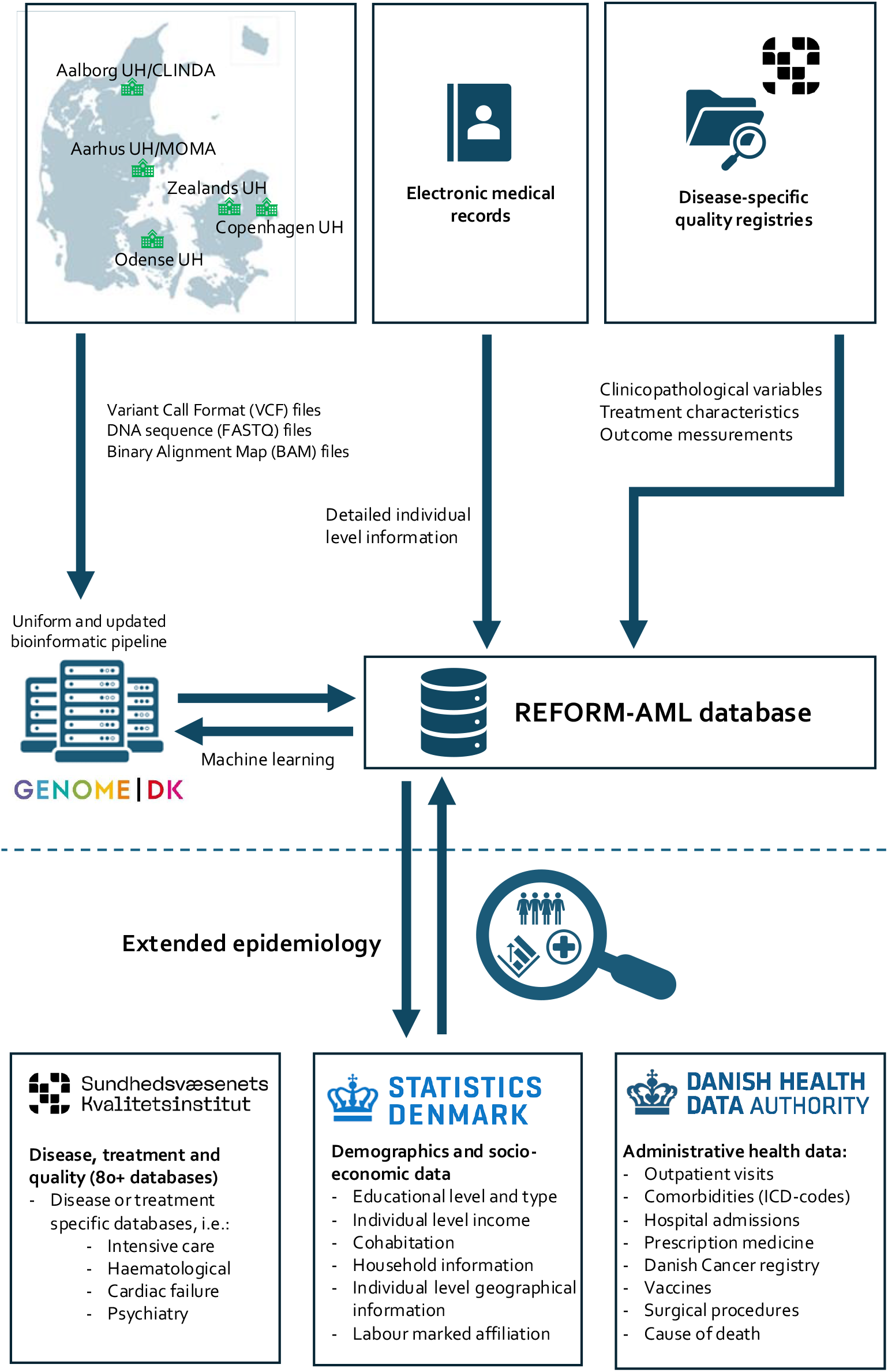
Dataflow and possibilities for external linkage of the REFORMAML database.

**Figure S2:**
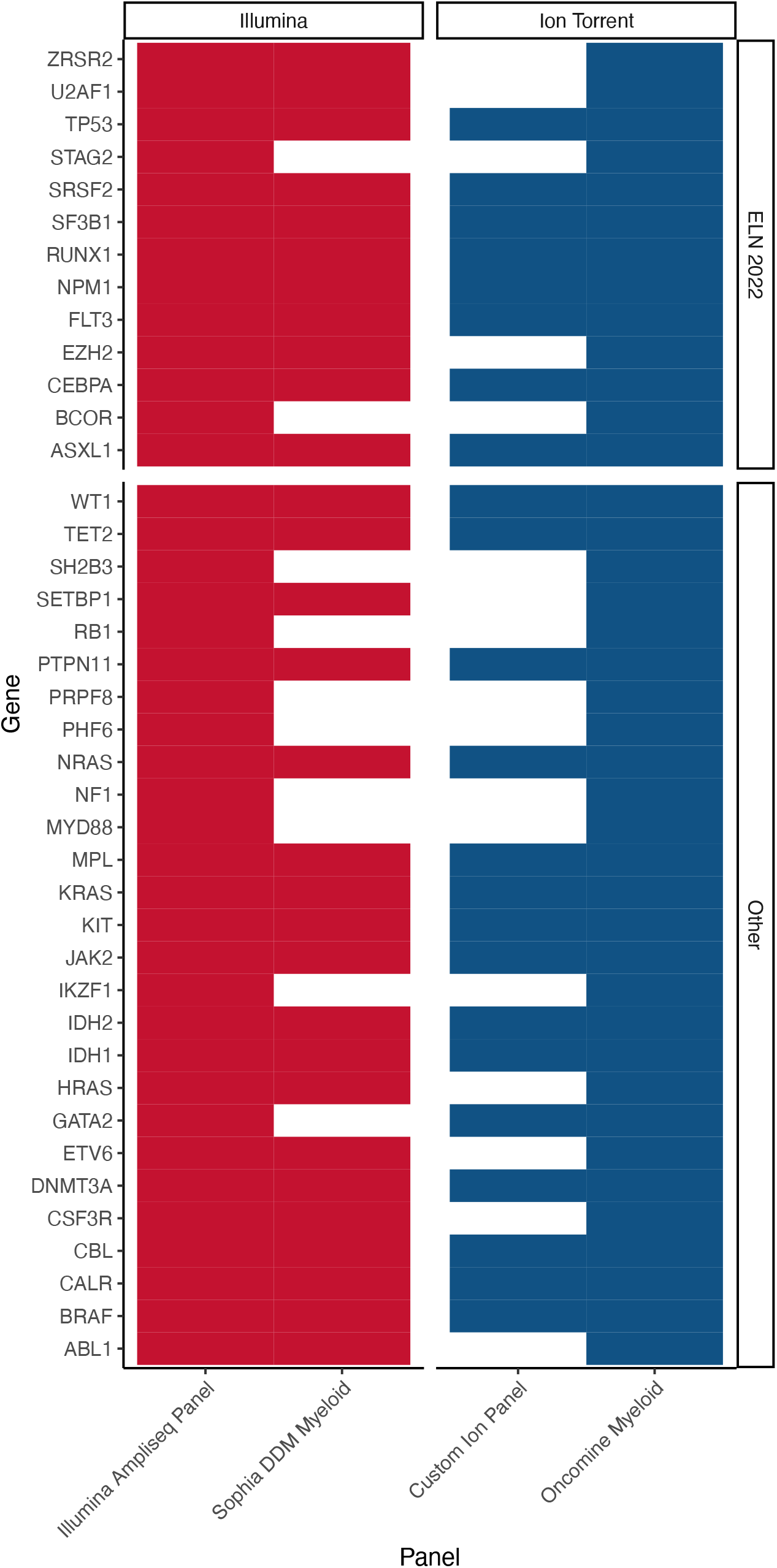
Next-generation sequencing panels used in the REFORM-AML dataset including covered genes. Numbers of patients for each panel: Illumina Ampliseq panel (n = 14), Sophia DDM myeloid (n = 322), Oncomine Myeloid (n = 275), and Custom Ion Panel (n = 174).

**Figure S3:**
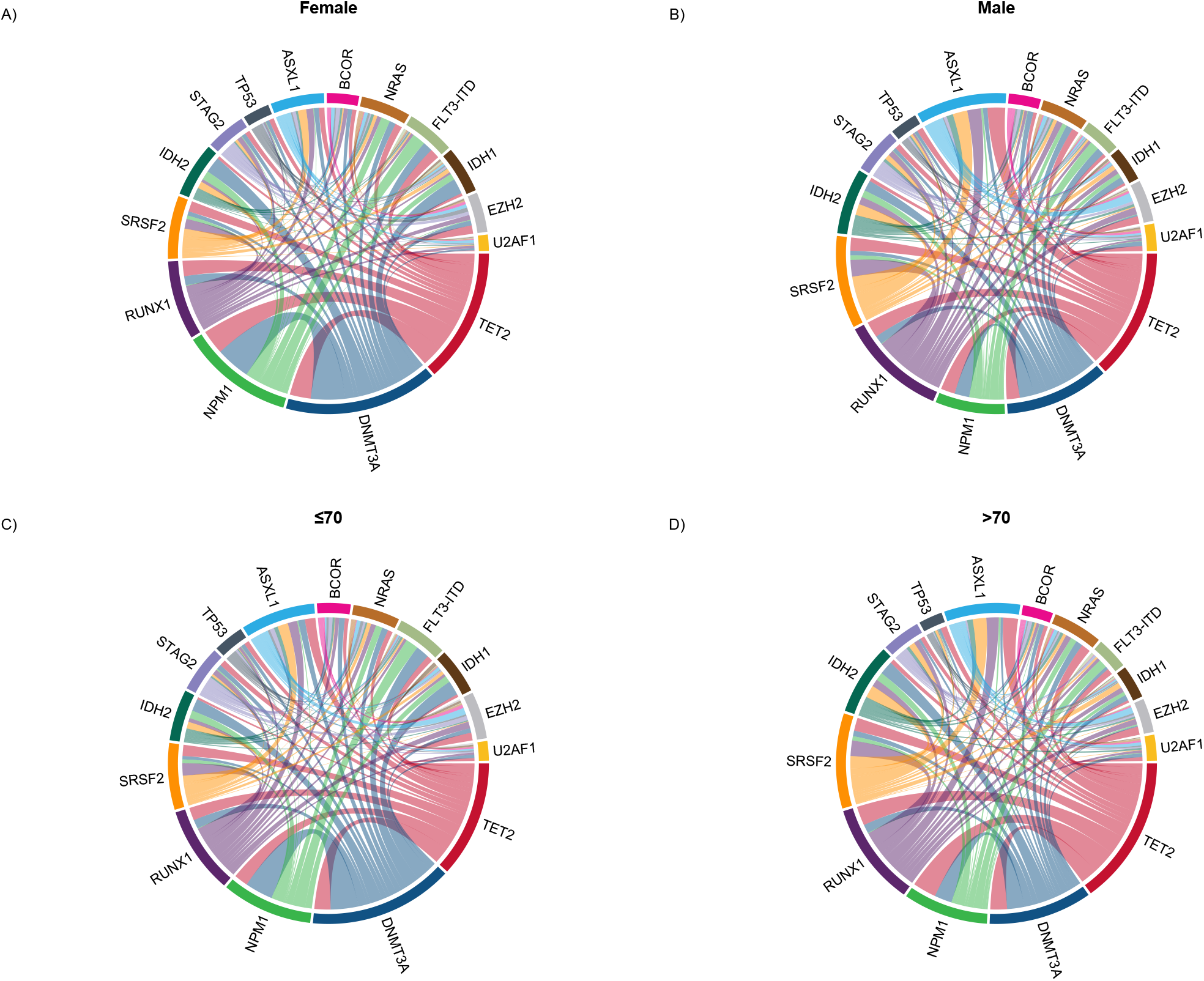
Circos plots showing the relative frequencies and pairwise co-occurrence of the 15 most commonly mutated genes in A) Female patients (*n* = 339), B) Male patients (*n* = 446), C) age ≤70 years (*n* = 390) and D) Age >70 years (*n* = 395).

## Supplementary tables

**Table S1:**
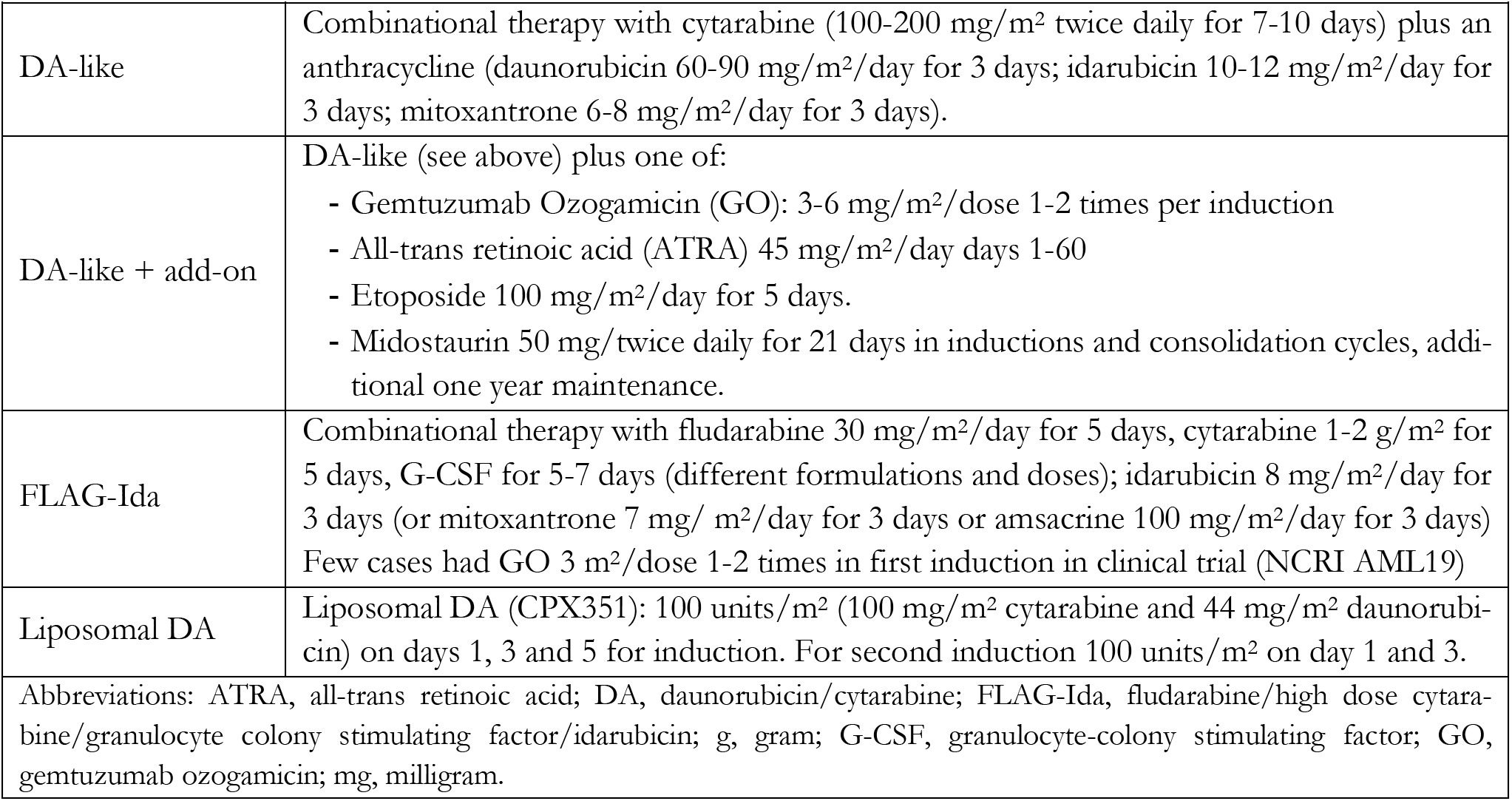
Treatment regimens for intensively treated cohort.

**Table S2:** Selected baseline characteristics of patients with AML and AML/MDS stratified on performed panel sequencing and age.

| Variable | All | ≤70 years (*n* = 1,013) |  |  | >70 years (*n* = 1,499) |  |  |
| --- | --- | --- | --- | --- | --- | --- | --- |
|  |  | NGS+ | NGS- | *p*-value | NGS+ | NGS- | *p*-value |
| **Numbers**, proportion of all (%) | 2,512 (100%) | 412 (16.4) | 601 (23.9) |  | 436 (17.4) | 1,063 (42.3) |  |
| **Sex**, male, *n* (%) | 1430 (56.9) | 219 (53.2) | 335 (55.7) | .455 | 265 (60.8) | 611 (57.5) | .263 |
| **Age**, median, years (IQR) | 73 (65-79) | 62 (52-67) | 62 (53-67) | .662 | 77 (73-81) | 79 (75-83) | <.001 |
| **Comorbidity index**, *n* (%) |  |  |  |  |  |  |  |
| ≤4 | 1,563 (62.2) | 370 (89.8) | 512 (85.2) | .075 | 209 (47.9) | 472 (44.4) | .245 |
| 5-8 | 818 (32.6) | 32 (7.8) | 73 (12.1) |  | 193 (44.3) | 520 (48.9) |  |
| ≥9 | 131 (5.2) | 10 (2.4) | 16 (2.7) |  | 34 (7.8) | 71 (6.7) |  |
| **Diagnostic period**, *n* (%) |  |  |  |  |  |  |  |
| 2015-2017 | 916 (36.5) | 57 (13.8) | 340 (56.6) | <.001 | 70 (16.1) | 449 (42.2) | <.001 |
| 2018-2020 | 1,010 (40.2) | 229 (55.6) | 166 (27.6) |  | 213 (48.9) | 402 (37.8) |  |
| 2021-2022 | 586 (23.3) | 126 (30.6) | 95 (15.8) |  | 153 (35.1) | 212 (19.9) |  |
| **Disease ontogeny\***, *n* (%) |  |  |  |  |  |  |  |
| *De novo* | 1,593 (63.4) | 296 (71.8) | 399 (66.4) | .179 | 278 (63.8) | 620 (58.3) | .138 |
| Antecedent hematological disease | 361 (14.4) | 38 (9.2) | 69 (11.5) |  | 69 (15.8) | 185 (17.4) |  |
| Therapy-related | 558 (22.2) | 78 (18.9) | 133 (22.1) |  | 89 (20.4) | 258 (24.3) |  |
| **BM blast**, median %, (IQR) | 40 (23-70) | 51 (27-76) | 50 (25-76) | .171 | 38 (22-61) | 34 (21-60) | .246 |
| **PB blast**, median %, (IQR) | 16 (2-46) | 19 (3-50) | 20 (3-53) | .870 | 9 (1-35) | 15 (2-43) | .002 |
| **Platelet count**, median, ×10<sup>9</sup>/L, (IQR) | 65 (33-120) | 66 (31-124) | 63 (33-114) | .884 | 79 (38-129) | 64 (32-115) | .008 |
| **WBC**, median, ×10<sup>9</sup>/L, (IQR) | 6.4 (2.1-32.1) | 8.0 (2.3-34.4) | 7.1 (2.1-35.3) | .821 | 4.4 (2.1-29.0) | 6.4 (2.1-31.1) | .257 |
| **LDH**, median, U/L (IQR) | 326 (216-605) | 374 (227-680) | 325 (212-656) | .101 | 281 (204-492) | 327 (217-602) | .002 |
| **Cytogenetics performed**, *n* (%) | 2,071 (82.4) | 390 (94.7) | 519 (86.4) | < .001 | 395 (90.6) | 767 (72.2) | <.001 |
| **Cytogenetic risk**<sup>#</sup>, *n* (%) |  |  |  |  |  |  |  |
| Favorable risk | 60 (2.9) | 21 (5.4) | 28 (5.4) | .995 | 4 (1.0) | 7 (0.9) | .953 |
| Intermediate risk | 1,520 (73.4) | 280 (71.8) | 374 (72.1) |  | 296 (74.9) | 570 (74.3) |  |
| Adverse risk | 491 (23.7) | 89 (22.8) | 117 (22.5) |  | 95 (24.1) | 190 (24.8) |  |
| **Treatment intensity**, *n* (%) |  |  |  |  |  |  |  |
| Intensive treatment | 906 (36.1) | 302 (73.3) | 420 (69.9) | .479 | 81 (18.6) | 103 (9.7) | <.001 |
| Low intensity | 782 (31.1) | 59 (14.3) | 94 (15.6) |  | 208 (47.7) | 421 (39.6) |  |
| Palliative/none | 824 (32.8) | 51 (12.4) | 87 (14.5) |  | 147 (33.7) | 539 (50.7) |  |
| **ASCT**, *n* (%) | 377 (15.0) | 145 (35.2) | 211 (35.1) | 1.0 | 11 (2.5) | 10 (0.9) | .034 |
| Abbreviations: BM, bone marrow; IQR, interquartile range (25th to 75th percentiles); LDH, lactate dehydrogenase; PB, peripheral blood; sAML, secondary AML; tAML, therapy related AML; WBC, white blood cell count; *Secondary disease takes precedence over therapy-related #Cytogenetic risk according to the European LeukemiaNet 2022 risk score[17]. |  |  |  |  |  |  |  |

